# State-wide Genomic Epidemiology Investigations of COVID-19 Infections in Healthcare Workers – Insights for Future Pandemic Preparedness

**DOI:** 10.1101/2021.09.08.21263057

**Authors:** Anne E. Watt, Norelle L. Sherry, Patiyan Andersson, Courtney R. Lane, Sandra Johnson, Mathilda Wilmot, Kristy Horan, Michelle Sait, Susan A. Ballard, Christina Crachi, Dianne J. Beck, Caroline Marshall, Marion Kainer, Rhonda Stuart, Christian McGrath, Jason C. Kwong, Pauline Bass, Peter G. Kelley, Amy Crowe, Stephen Guy, Nenad Macesic, Karen Smith, Deborah A. Williamson, Torsten Seemann, Benjamin P. Howden

## Abstract

**Background:** COVID-19 has resulted in many infections in healthcare workers (HCWs) globally. We performed state-wide SARS-CoV-2 genomic epidemiological investigations to identify HCW transmission dynamics and provide recommendations to optimise healthcare system preparedness for future outbreaks.

**Methods:** Genome sequencing was attempted on all COVID-19 cases in Victoria, Australia. We combined genomic and epidemiologic data to investigate the source of HCW infections across multiple healthcare facilities (HCFs) in the state. Phylogenetic analysis and fine-scale hierarchical clustering were performed for the entire Victorian dataset including community and healthcare cases. Facilities provided standardised epidemiological data and putative transmission links.

**Findings:** Between March and October 2020, approximately 1,240 HCW COVID-19 infection cases were identified; 765 are included here. Genomic sequencing was successful for 612 (80%) cases. Thirty-six investigations were undertaken across 12 HCFs. Genomic analysis revealed that multiple introductions of COVID-19 into facilities (31/36) were more common than single introductions (5/36). Major contributors to HCW acquisitions included mobility of staff and patients between wards and facilities, and characteristics and behaviours of individual patients including super-spreading events. Key limitations at the HCF level were identified.

**Interpretation:** Genomic epidemiological analyses enhanced understanding of HCW infections, revealing unsuspected clusters and transmission networks. Combined analysis of all HCWs and patients in a HCF should be conducted, supported by high rates of sequencing coverage for all cases in the population. Established systems for integrated genomic epidemiological investigations in healthcare settings will improve HCW safety in future pandemics.

**Funding:** The Victorian Government, the National Health and Medical Research Council Australia, and the Medical Research Future Fund.

## Introduction

The COVID-19 pandemic has resulted in the hospitalization of large numbers of patients with severe disease, particularly in older age groups. ^1^ Healthcare workers (HCWs) on the frontline have acquired COVID-19 in many different settings, often despite adequate availability and choice of appropriate personal protective equipment (PPE).^2-6^ To optimise the safety of HCWs and patients, it is critical for hospital infection control teams and, more broadly, healthcare systems to understand the drivers of infections in HCWs, through systematic investigations of the circumstances around these putative transmissions in healthcare settings. Internationally, genomics of SARS-CoV-2 has been a powerful tool for understanding transmission links and outbreaks.^7-10^ Whilst the investigation of HCW infections has traditionally been achieved through epidemiologic assessments, combined genomic and epidemiologic analyses have now emerging as the new standard-of care for these investigations.^11,12^

The state of Victoria, Australia (population ∼6.7 million)^13^ experienced two waves of COVID-19 in 2020. Comprehensive prospective genomic sequencing of SARS-CoV-2-positive samples was undertaken by the public health genomic reference laboratory (the Microbiological Diagnostic Unit – Public Health Laboratory (MDU-PHL)), with samples sequenced from 75% of cases. The first wave was largely a polyclonal outbreak, characterised by multiple introductions from overseas travellers with limited onwards transmission in the population, and very limited transmission to HCWs.^6,14^ The second wave in Victoria was largely a clonal outbreak, centred in Melbourne, Victoria, originating from a breach in the hotel quarantine system for returned travellers.^7^ This second wave resulted in outbreaks occurring across many healthcare facilities (HCF) and aged care facilities (ACF).^7^ Globally, HCWs are at increased risk of infection with coronavirus disease (COVID-19).^2^ Multiple studies are beginning to document nosocomial transmission and infection in HCWs^11-12,15-16^ and highlighting the need for tailored infection control investigations and responses. Whole genome sequencing can contribute high resolution data to describe and investigate such transmission networks.

Here we describe the process and findings of investigations of HCW infections in multiple HCFs across our state. We hypothesised that an integrated genomic epidemiological analysis of COVID-19 HCW infections, interpreted in the broader context of all healthcare and community infections, would enhance understanding of the source of HCW infections and identify common transmission risks. Our results aim to provide a framework for workflows and metadata required to maximise HCF preparedness to investigate COVID-19 HCW infections, and optimise staff safety for future outbreaks.

## Methods

### Setting and data sources

This project was undertaken in the state of Victoria, Australia (population ∼6.7 million),^13^ where the healthcare network includes eleven major metropolitan health services. Since the start of the COVID-19 pandemic, all samples positive for SARS-CoV-2 by RT-PCR are requested to be forwarded to MDU-PHL for genomic sequencing.^7,14,17^ Prospective sequencing was conducted on all samples received at MDU-PHL, with samples sequenced from approximately 75% of cases.^7^

The genomic epidemiology team at MDU-PHL assisted all HCFs requesting genomic investigations of COVID-19 outbreaks in HCWs (and often including patients) in their facilities. Investigations were conducted to inform operational improvements at each healthcare facility, including infection prevention and control, for infection control purposes, with each healthcare facility providing the epidemiological data to inform the genomic epidemiological investigation. Investigations were an iterative process developed through collaboration with healthcare facilities, refined to a standard workflow and list of required and desirable metadata (Box. 1). Some of these investigations were conducted in near to real time whilst others were requested retrospectively once capacity was available at the HCF to perform the epidemiological assessment. For this study, HCWs were defined as any staff, students or volunteers working in a hospital or paramedic setting, excluding community residential aged care facilities (RACFs).

### Genomic data and bioinformatic analysis

Detailed methods are described elsewhere;^7,14^ briefly, extracted RNA from SARS-CoV-2 RT-PCR positive samples underwent tiled amplicon PCR using either ARTIC version 1 or 3 primers,^18^ following published protocols.^19^ Reads were aligned to the reference genome (Wuhan Hu-1; GenBank MN908947.3) and consensus sequences generated. Quality control (QC) metrics on consensus sequences included requiring ≥65% genome recovered, ≤35 single nucleotide polymorphisms (SNPs) from the reference genome, and ≤300 ambiguous or missing bases. A single sequence was selected from each patient for phylogenetic analysis. Genomic clusters were defined as two or more related sequences using a complete-linkage hierarchical clustering algorithm of pairwise genetic distances derived from a maximum likelihood phylogenetic tree. Genomic clustering was used to identify plausible genomic links between cases, which were further interpreted together with epidemiological data.

### Combined genomic and epidemiologic analysis

Genomic epidemiological analyses were performed in three stages (**Box 1**). Beginning with a line list from HCFs identifying HCW and patients with sufficient identifiers to match to available lab and genomic data. Stage one linked cases with samples and grouped cases by genomic cluster, identifying the minimum number of genomic introductions likely to have taken place, and formed the foundation for all further investigations. Stage two expanded the investigation by including the case information such as date of sample collection, symptom onset and diagnosis for each individual. The results of this step allowed for focusing of further epidemiological investigations. Stage three provided in-depth epidemiological investigation of genomic clusters by combining epidemiological location and exposure data.

Results of each analysis were reported to the facilities as an iterative process, with collaborative meetings cases included in the analysis were reviewed, then the genomic data were presented. Facilities were given the opportunity to review and add any epidemiological data to assist with the analysis and to put forward any specific queries based on their epidemiological analysis. The analyses were then refined based on the outcomes of the meetings and compiled into a final report.

### Ethics

Ethical approval was received from the University of Melbourne Human Research Ethics Committee (study number 1954615.4).

## Results

Between March and October, 2020, MDU-PHL were approached by 12 HCFs to assist with genomic epidemiological investigations into HCW COVID-19 cases. Investigations ranged in scope from individual suspected transmission events to ward- or facility-level investigations. MDU-PHL assisted with 36 investigations, with 9/12 facilities requesting more than one investigation. The majority of investigations were undertaken in large public university hospitals, with a small number of private facilities, including a total of 21 campuses and more than 9900 beds,^20^ as well as the metropolitan paramedic service. A total of 765 HCWs and 1,273 patients were investigated, with sequencing available for 80% (612) of HCWs and 80.8% (1,028) of patients (data summarized in **Table 1)**.

**Table 1.**
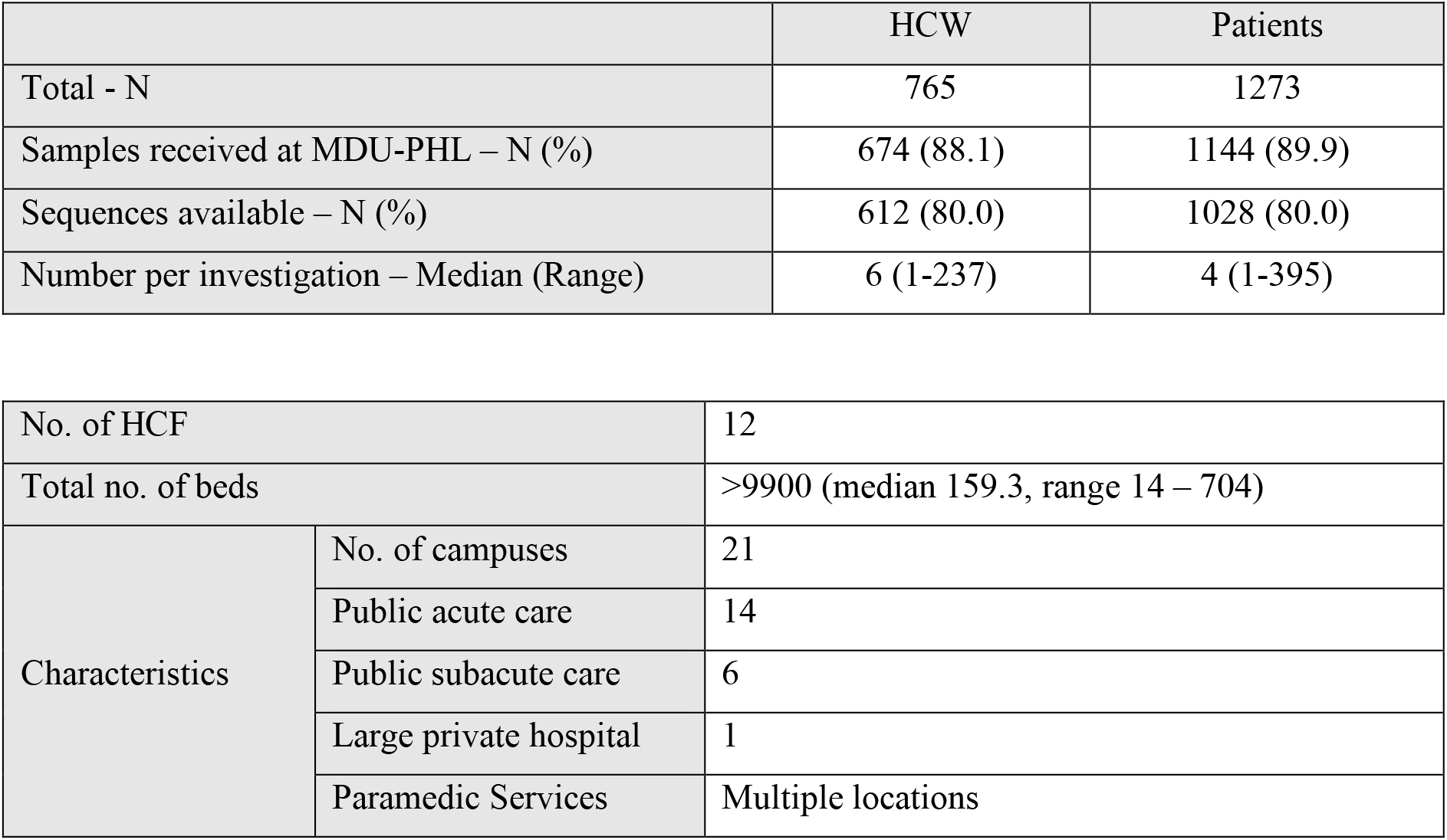
Summary of the 36 genomic epidemiological investigations

For the five healthcare networks where we performed analyses across the whole institution (all campuses), an estimated 59% to 80% of HCW infections were deemed to be likely acquired at the HCF.

### Genomic results often, but not always, had high concordance with epidemiologic investigations

Genomic analysis provides an estimation of the minimum number of introductions to a facility through the number of genomic clusters present. The median number of introductions in these analyses was 3 per facility (IQR 2 - 8, range 1 – 35) and the median number of HCWs per genomic cluster was 1 (IQR 1-3, range 1 – 104). These analyses found that 31/36 (86.1%) investigations included cases resulting from multiple introductions, while 5/36 (13.9%) investigations involved a single introduction. Investigations with multiple introductions had a median of 6 HCWs (IQR 1 – 17, range 1 - 237) and 7 patients (IQR 3 – 39 range 1 - 395) while investigations with single introductions had a median of 1 HCW (IQR 1 – 6, range 1 – 7) and 2 patients (IQR 1 – 36, range 1 – 56). Thirteen of these analyses were instances of investigations into single staff members and their contacts; three of these could not be resolved as sequence data for the case or contacts was unavailable. While it is more likely to have multiple genomic introductions when there are high case numbers present at a facility, we found that low case numbers did not always result in fewer genomic introductions.

In these investigations, we largely observed high levels of concordance between epidemiological hypotheses (healthcare acquired infection or not) and genomic data where transmission had occurred, with some notable exceptions. One, a multi-campus facility, epidemiologically identified multiple individual outbreaks within their campuses. The combined genomic epidemiological analysis found undetected transmission events and that most of the individual outbreaks and unlinked cases were linked back to a single introduction or source (**Figure 1, A**). Conversely, Facility B experienced a large outbreak at one campus; genomics identified three concurrent outbreaks from separate genomic clusters at a time of high community prevalence (**Figure 1, B**). In both cases, genomic data significantly altered the understanding of transmission in the facilities, leading to changes in infection control practices. For example, at one HCF, upon reviewing the epidemiology in the light of the genomic data it become clear that some epidemiological links were missed, highlighting the need to strengthen contact tracing applications and resources for this facility.

**Figure 1.**
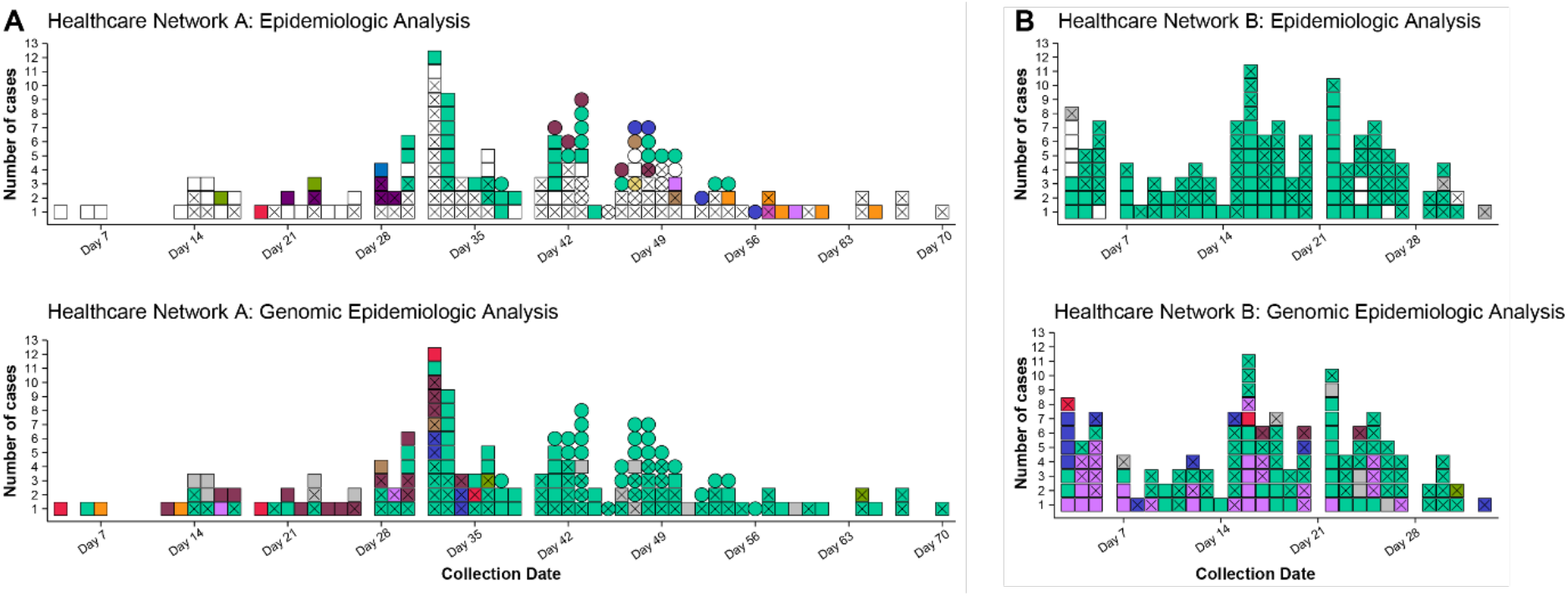
Comparison of clustering (identifying cases of HCW and patient infections that are likely to be related) by epidemiology and genomics analyses at two facilities. Colour indicates cluster (epidemiological cluster for epidemiologic analyses and genomic cluster for genomic epidemiological analyses); white indicates unknown cluster/acquisition; grey indicates non-healthcare acquired infection; X indicates HCW case; squares and circles in panel A indicate two different campuses of the healthcare network. ‘Collection date’ refers to day of the outbreak period for each panel. **Panel A**. Epidemiological analysis of COVID-19 cases at Facility A (two separate campuses) identified 12 epidemiologic clusters of likely transmission and 88 cases with no known acquisition source. Genomic epidemiologic analysis for the same network showed that the vast majority of cases were linked within eight genomic clusters, including one dominant cluster (lighter green), and only 12 cases not genomically linked to the HCF. **Panel B**. Epidemiological analysis of cases at Facility B identified >100 HCW cases likely acquired at the facility, all thought to be part of a single epidemiologic cluster, and nine HCW cases not thought to be healthcare acquired. Genomic epidemologic analysis indicated multiple introductions, rather than a single introduction, with six different genomic clusters co-occurring, and only six cases not genomically-linked to the HCF.

### Mobility of HCWs and patients often implicated in hospital transmissions

A common theme from HCFs was that many infections resulted from the mobility of staff or patients. Movement of staff and patients between wards and campuses while pre-symptomatic or asymptomatic was implicated in dissemination of COVID-19 between facilities within hospital networks (4/8 facilities where multiple campuses were investigated). At one facility, a single patient was found to have seeded cases in two wards due to transport while asymptomatic. Their movement between general and rehabilitation wards resulted in spread to 5 naïve patients and 15 HCWs. Identification of spread due to patient mobility led to one HCF to introduce asymptomatic testing for any patient moving from acute to subacute ward during periods of high community transmission.

### Patient features or behaviours contributing to COVID-19 transmission to HCWs

In the course of these investigations, elderly patients with altered mental states were found to exhibit behaviours that contributed to the spread of COVID-19 within at least four HCFs. Patients suffering from delirium or dementia were often highly mobile (wandering behaviours) and exhibiting aerosol-generating behaviours (coughing, shouting or singing). Due to the nature of these patients and their increased need of HCW support, direct contact was often implicated in the transmission. In these cases, combined genomic and epidemiological data showed that one or more patients, admitted from a single ACF at the same time, were found to be the likely acquisition source for staff that contracted COVID-19 working on a ward for COVID-19 positive patients with dementia or delirium.

### Limiting the scope of investigations may lead to erroneous conclusions

Investigations limited to a single ward were found to have limited utility when performed at large facilities with high numbers of positive cases. These investigations often found cases without any known transmission source, transmission, with multiple outbreaks deemed separate by epidemiological investigations, subsequently identified as single outbreaks by genomics. For example, investigation of a ward-based outbreak at one facility identified eight genomically-linked cases. An expanded investigation, including all HCWs and patients at the facility in a similar time, identified an additional 10 cases were part of the same genomic transmission network as the first ward, indicating that cryptic transmission had likely occurred from the first ward analysed, and providing opportunities for further targeted epidemiologic investigations (**Figure 2**).

**Fig 2.**
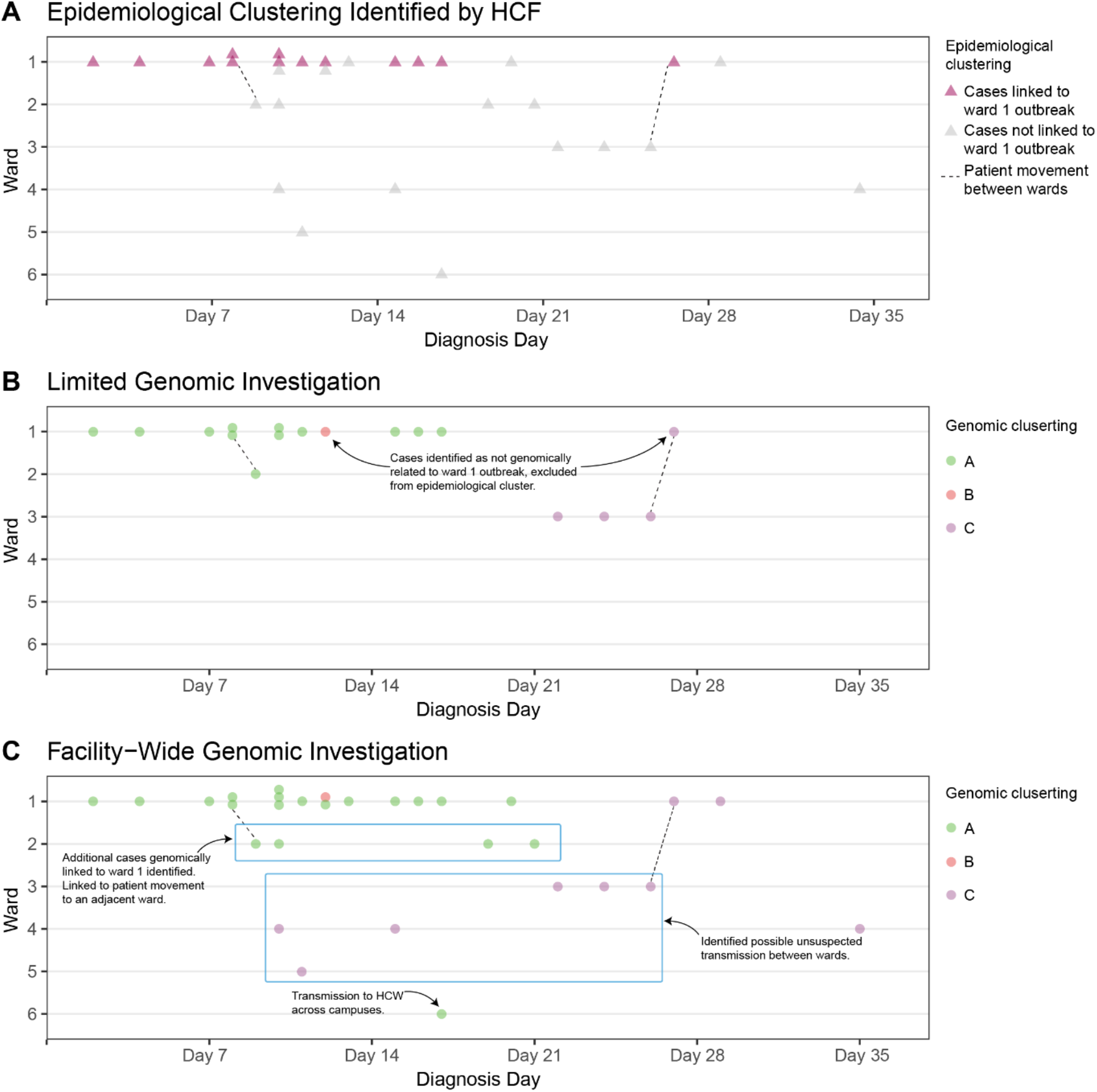
Comparison of clustering (identifying cases of HCW and patient infections that are likely to be related) at one multi-campus healthcare facility using three different models: epidemiological clustering (panel A), limited genomic investigation (cases in a single ward selected by HCF, panel B), and facility-wide genomic infections (panel C). Each panel shows the distribution of cases (triangles in panel A, circles in panels B and C) across six different wards (Wards 1-6) over a six-week time period in 2020, where ‘Diagnosis day’ refers to the day in the outbreak period. In **panel A**, thirteen cases were identified by the HCF as a likely epidemiologic cluster (pink triangles). These cases, with the addition three cases from adjacent ward (Ward 3) were submitted for a limited genomic investigation (**panel B**); cases (circles) are coloured by genomic cluster. This showed that most of the cases submitted were part of the same genomic cluster, but two of the Ward 1 cases were not linked (one case from GC B, and one case from GC C, which was linked to two other cases on Ward 3). **Panel C** shows a broader facility-wide genomic investigation that was undertaken to investigate cases on other wards; all HCW and patient cases were included in the facility-wide investigation. This genomic analysis found the main outbreak from Ward 1 was larger than first identified, linking outbreaks in adjacent wards to the Ward 1 outbreak, with cryptic transmission between wards resulting in spread, including transmission to another hospital campus. Unexpected links were also identified for GC C, with cases spread over four wards. These genomic links were used to direct further investigations to identify causes of transmission and introduce mitigation strategies.

Similarly, limitations were found when examining investigations without contemporaneous genomic data from community cases. Lack of sufficient community cases for context can lead to inaccurate interpretation of transmission events. Initial investigations performed for one Facility C without community context indicated likely transmission between HCWs in a work setting. The same data, when interpreted with community context, indicated that transmission was more likely to have occurred in a social setting external to the workplace, and confirmed by further epidemiologic investigations (**Figure 3**).

**Fig 3.**
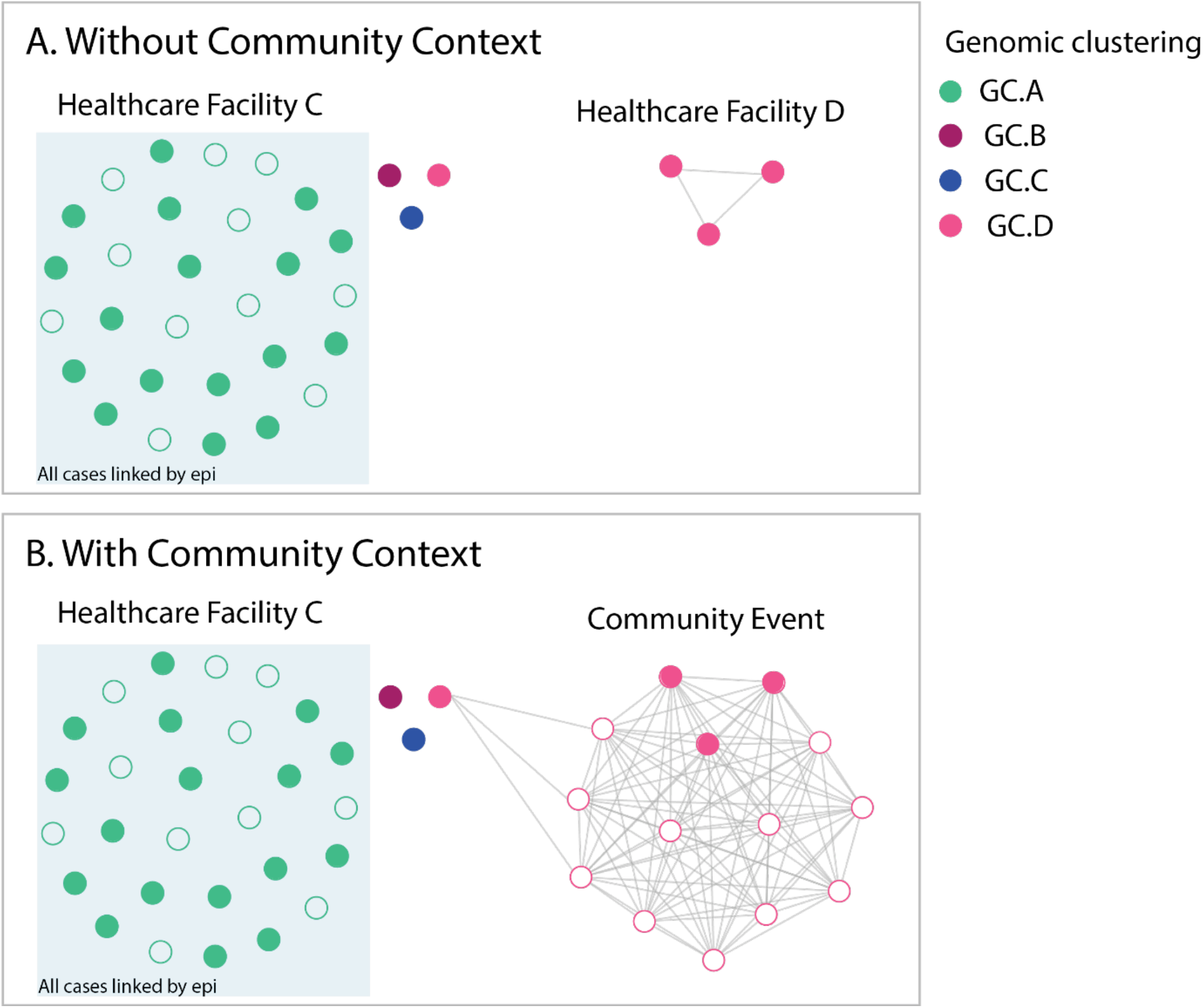
Comparison of genomic epidemiological analyses analysed with and without genomic data for community cases. Filled circles indicate HCWs, unfilled circles indicate non HCWs, colour indicates genomic cluster. **Panel A** shows analysis of cases from facility C (mostly linked by epidemiology and genomics with dominant genomic cluster GC A (green), and three additional HCW cases from different genomic clusters (genomic clusters GC B, GC C and GC D), plus three cases at facility D (related to each other) from genomic cluster GC D. In isolation, this suggests possible cryptic transmission between the two healthcare facilities. Addition of community sequences into the analysis (**Panel B**) demonstrated that the HCWs at both facility C and facility D likely acquired infection from a social event in the community that was attended by these cases.

### Key learnings for genomic investigations of HCW infections

The collaborative meetings with HCFs provided an opportunity to educate clinicians about the utility and limitations of genomic analyses, share initial findings from the genomic analysis, add additional relevant epidemiological data to assist with interpretation, gauge the understanding of the genomic results and clarify further where necessary. They also provided an opportunity for additional epidemiological data that may have been missed during data collection, such as data on social links between cases e.g., staff often socialised together after working hours or lived in shared housing with other HCWs that maybe from the same or other HCFs, which is difficult to capture in standard line lists shared as part of the early investigation process. Anecdotally, one HCF identified that 50% of their HCWs lived with other HCWs.

## Discussion

The COVID-19 pandemic has reinforced the need to optimise HCF systems to protect both patients and HCWs from infectious diseases threats.^21^ Here we detail genomic epidemiological investigations undertaken by a reference public health genomics laboratory of COVID-19 infections in HCWs across multiple facilities in Victoria, Australia and define a framework for this type of activity in future. Through an iterative, collaborative process with 12 HCFs, we performed 36 investigations for 765 HCWs out of a total of 1240 HCW infections notified for the state.^22^ Underpinning these analyses was efficient case ascertainment and a very high proportion of positive cases sequenced, including samples from HCWs and patients as well as the community. Several of these investigations were conducted in near to real time which allowed facilities to rapidly change infection prevent protocols to limit further spread. A clear strength of the investigative process in this study was establishing a forum of laboratory and clinical experts to initiate, discuss and progress investigations which facilitate the integration of genomic results with infection prevention and control methods.

This study highlights important commonalities that were seen across the facilities investigated and the importance of understanding SARS-CoV-2 transmission for future outbreak prevention. We found that physical movement of individuals as well as aerosol generating behaviours led to known and cryptic transmission of COVID-19 within the facilities we investigated. While this pattern of staff and patient movement is likely ubiquitous to HCFs and has been seen to contribute to the spread of COVID-19 elsewhere,^11,12^ it highlights the importance of investigating all positive cases of HCWs and patients within a facility. We noted instances such as at Facility A, where the genomic data refuted the findings of the epidemiologic data, interpretation of the two data sets together would significantly change the infection control response. Similar scenarios were found by Meijer et al.^23^

While genomic analyses can be informative with basic epidemiological data, the rich detail added by comprehensive epidemiological data dramatically improves their utility. Rapid and effective data capture and management was a significant challenge for most facilities during the epidemic, delaying and limiting infection control investigations; implementation of sustainable continuous data collection processes within HCFs should be a priority for future epidemic preparedness, allowing earlier initiation of epidemiological and genomic investigations.

Based on our experiences, we propose a set of minimum and enhanced metadata and a workflow to optimise the utility of HCW investigations (**Box 1**), recognising that expansion and resourcing for such systems can vary between facilities. Wherever possible, integration with existing data systems should be leveraged, such as data from employee databases. Metadata should be collected in standardized templates, and captured in a secure version-controlled database (e.g. REDCap). This maintains data integrity during staff turnover or when surge capacity is called for in response to events. The World Health Organisation (WHO) has outlined the minimum metadata to ensure that genomic sequencing of SARS-CoV-2 samples will be of most use.^24^ From our experiences here, we propose that these metadata should ideally be expanded when performing genomic epidemiological analysis. To allow for rapid utilisation of data when the need arises, prior consideration should be given to the governance framework for the use and integration of the data into other systems, such as disclosure to public health laboratories during investigations, and its relationship to data captured by other public health organisations.

Limitations of the study include the highly clonal nature of cases in Victoria at this time, with >95% of cases from the second wave being seeded for a single tm event. This limited the ability to resolve some transmission networks, particularly early in the outbreak, and may erroneously suggest single introductions of a cluster when there may have been multiple introductions from a genomic cluster from the community. This increases the importance of quality epidemiologic data to assist with interpretation of genomic data when performing these analyses. Our investigations were also limited by HCW and patient cases that were not able to be sequenced although numbers were relatively small, and the proportion of cases successfully sequenced was greater than most other jurisdictions. Similar processes could easily be applied to other healthcare systems where genomics is less commonly available; in particular, focussed sequencing of hospitalised cases and HCWs could achieve very similar results, albeit with a small chance of false-positive genomic links due to multiple introductions of the same genomic cluster from the community.

The results from each facility have shown that there were multiple contributors to COVID-19 infections in HCWs in Victoria in 2020, and that while there were common factors contributing to transmission across different facilities, each outbreak was in fact a unique combination of contributors and had to be assessed individually. Through our experience working with multiple HCFs, we found that it was essential to investigate all positive HCW and patient cases in a facility along with detailed epidemiological data, wherever feasible. Collaborative and interactive exploration of the combined data uncovered further epidemiological links, maximising the impact of the analyses for the HCF, and providing the greatest opportunities for HCFs to optimise the safety of HCWs and patients in the future.

## Supporting information

STROME-ID checklist

## Data Availability

All consensus sequences and Illumina sequencing reads are available at https://github.com/MDU-PHL/COVID19-paper. Sequence accession numbers can be found in Supplementary Table 1 (note: some cases were included in investigations at more than one facility).

## Data sharing

All consensus sequences and Illumina sequencing reads are available at https://github.com/MDU-PHL/COVID19-paper.

## Acknowledgements

We gratefully acknowledge the large number of staff at Victorian healthcare facilities and diagnostic laboratories who collected data and undertook diagnostic testing for COVID-19 in Victoria. We would particularly like to acknowledge the considerable efforts of infection prevention and control staff at affected facilities. We would especially acknowledge the contribution of Terri Butcher and Adrian Tramontana (Western Health), Andrew Stewardson, Amanda Dennison, Adam Jenny, Anton Peleg, and Denis Spelman (Alfred Health), Paul Van Buynder and Emily Tait (Ambulance Victoria), Claire Gordon (Austin Health), Kylie Hall and Alexandra Bonello (Eastern Health), Bradley Gardiner and Suman Majumdar (Epworth), Tony Korman and Maryza Graham (Monash Health), Victoria Madigan, Barbara Brozic, Madelaine Flynn, and Craig Aboltins (Northern Hospital), Susan Gonelli and Srikanth Velandai (Peninsula Health), Katherine Bond, Vivian Leung, Chris Bailie, Laura Piu, Jackie O’Connor, and Kirsty Buising (Royal Melbourne Hospital), Leilani Knapp, Mary-Jo Waters, Yves Lorenzo, Lydia Sims, and Samantha Palmby (St Vincent’s Hospital).

## BOX 1

**Genomic Epidemiological Investigations**

### Part 1 – Establishing basic genomic and epidemiologic data

1. Establish HCF transmission hypotheses for investigation
2. Collect case list and metadata (demographic & case information).
3. Identify missing data, follow up on sample and sequencing availability.
4. Build phylogenetic tress with suitable context isolates (temporal & geographic).
5. Match metadata to available genomic data.
6. Discuss genomic clustering with HCF.
  a. Optional stopping point

### Part 2 – Integrating case information

7. Overlay detailed epidemiological metadata (date of diagnosis, and patient/staff role)
8. Discuss with HCF the concordance between epidemiological data and phylogenetic data.

### Part 3 – Integrating exposure and location data

9. Overlay detailed epidemiological location data & exposure data (known exposure events)
10. Refine genomic clustering with detailed epidemiological metadata.
11. Final written report.

#### Optimal metadata to include

##### Individual level metadata

1. Demographic data
  a. Name
  b. Date of birth
  c. Lab / UR number
2. Case information
  a. Date of diagnosis, Date of onset, Date of collection
  b. Role - HCW (with or without patient contact; specific role) / Patient / Visitor
3. Location data
  a. Patient admission date, ward and bed number and movement details
  b. Staff shift dates, primary and secondary locations (where available)
  c. Furlough
4. Exposure data
  a. Known COVID positive contacts with dates of contact
  b. PPE breach or other known high-risk events – positive cases, contact level
  c. Staff links to other HCF or ACF
  d. Travel History international and local
  e. Contact with other staff outside the workplace e.g. car-pooling or social events Staff living with / links to other HCW ACW
  f. Residence in or exposure to community “hotspot” (a location of intense community transmission)

##### Facility level metadata

a. PPE donning and doffing procedures /locations
b. Staff facilities, e.g. shared team rooms
c. Facility links to other HCF or ACF

