## Supplementary material for "State-wide Genomic Epidemiology Investigations of COVID-19 Infections in Healthcare Workers – Insights for Future Pandemic Preparedness": STROME-ID checklist

| **Title and abstract** | **Item** | **STROBE-ID items** | **STROME-ID items** | **Responses** |
| --- | --- | --- | --- | --- |
| Introduction | 1 | (a) Indicate the study’s design with a commonly used term in the title or the abstract (b) Provide in the abstract an informative and balanced summary of what was done and what was found | STROME-ID 1.1: the term molecular epidemiology should be applied to the study in the title or abstract and the keywords when molecular and epidemiological methods contribute substantially to the study | ‘Genomic epidemiology’ included in title |
| Background rationale | 2 | Explain the scientiﬁc background and rationale for the investigation being reported | STROME-ID 2.1: provide background information about the pathogen population and the distribution of pathogen strains within the host population at risk | Explained in introduction and methods  Population – SARS-CoV-2 PCR positive patients and healthcare workers (HCWs), as identified by healthcare facilities (HCFs), with genomic sequencing results available (% sequenced for each group included in results) |
| Objectives | 3 | State speciﬁc objectives, including any prespeciﬁed hypotheses | STROME-ID 3.1: state the epidemiological objectives of using molecular typing | Stated in last paragraph of introduction – to understand source of HCW infections and identify common transmission risks |
| **Methods** |  |  |  |  |
| Study design | 4 | Present key elements of study design early in the paper | .. |  |
| Molecular terminology |  | .. | STROME-ID 4.1: deﬁne or cite deﬁnitions for key molecular terms used within the study (eg, strain, isolate, and clone) | Definitions for genomic clustering included in methods |
| Molecular markers |  | .. | STROME-ID 4.2: clearly deﬁne the molecular markers that were used with a standard nomenclature | N/A |
| Infectious disease case deﬁnition |  | .. | STROME-ID 4.3: clearly state the infectious-disease case deﬁnitions | Stated in methods (SARS-CoV-2 PCR positive) |
| Laboratory methodology |  | .. | STROME-ID 4.4: describe sample collection and laboratory methods, including any methods used to minimise and measure cross-contamination, and give the criteria used to interpret strain classiﬁcation | Detailed in methods  Cross-contamination not addressed as clinical samples (i.e. addressed using usual diagnostic microbiology lab methods) |
| Setting | 5 | Describe the setting, locations, and relevant dates, including periods of recruitment, exposure, follow-up, and data collection | STROME-ID 5.1: clearly state the timeframe of the study; consider and appropriately reference the molecular clock of markers if known, and the natural history of the infection | Timeframe March to October 2020 |
| Participants | 6 | (a) *Cohort study*—give the eligibility criteria, and the sources and methods of selection of participants. Describe methods of follow-up *Case-control study*—give the eligibility criteria, and the sources and methods of case ascertainment and control selection. Give the rationale for the choice of cases and controls   *Cross-sectional study*—give the eligibility criteria, and the sources and methods of selection of participants   (b) *Cohort study*—for matched studies, give matching criteria and number of exposed and unexposed   *Case-control study*—for matched studies, give matching criteria and the number of controls per case | STROME-ID 6.1: state the source of participants and clinical specimens, and clearly describe sampling frame and strategy | Observational study  Stated in methods – HCFs identified HCWs and patients for analysis based on their epidemiologic investigations |
| Variables | 7 | Clearly deﬁne all outcomes, exposures, predictors, potential confounders, and eﬀect modiﬁers. Give diagnostic criteria, if applicable |  | Not directly relevant (observational) |
| Data sources/ measurement | 8* | For each variable of interest give sources of data and details of methods of assessment (measurement). Describe comparability of assessment methods if there is more than one group |  | Not directly relevant (observational) |
| Multiple-strain infections |  | .. | STROME-ID 8.1: describe any methods used to detect multiple-strain infections and measure their eﬀect on the study ﬁndings | Not addressed in this study as this is not routine practice for SARS-CoV-2 genomics |
| Bias | 9 | Describe any eﬀorts to address potential sources of bias | STROME-ID 9.1: describe any eﬀorts made to address discovery or ascertainment bias | Observational study, dependent upon availability of sequencing. Attempted to address by analysing data at a whole facility level for several HCFs to illustrate potential bias/incomplete results when only analysing a subset of cases based on epi assessment (Figure 2) |
| Study size | 10 | Explain how the study size was arrived at | STROME-ID 10.1: describe any unique restrictions placed on the study sample size | Observational |
| Quantitative variables | 11 | Explain how quantitative variables were handled in the analyses. If applicable, describe which groupings were chosen, and why | ·· | N/A |
| Statistical methods | 12 | (a) Describe all statistical methods, including those used to control for confounding (b) Describe any methods used to examine subgroups and interactions (c) Explain how missing data were addressed (d) *Cohort study*—if applicable, explain how loss to follow-up was addressed *Case-control study*—if applicable, explain how matching of cases and controls was addressed   *Cross-sectional study*—if applicable, describe analytical methods taking account of sampling strategy  Describe any sensitivity analyses | STROME-ID 12.1: state how the study took account of the non-independence of sample data, if appropriate  STROME-ID 12.2: state how the study dealt with missing data | Only median/IQR/range used in this paper |
| **Results** |  |  |  |  |
| Participants | 13* | (a) Report the numbers of individuals at each stage of the study (eg, numbers potentially eligible, examined for eligibility, conﬁrmed eligible, included in the study, completing follow-up, and analysed) (b) Give reasons for non-participation at each stage   Consider use of a ﬂow diagram | STROME-ID 13.1: Report numbers of participants and samples at each stage of the study, including the number of samples obtained, the number typed, and the number yielding data | Participant numbers (potential and included) described in results, and background of total number of HCW cases and community cases of COVID-19 in Victoria also described in results. Cases submitted for sequencing and passing QC also included in results. |
|  |  |  | STROME-ID 13.2: if the study investigates groups of genetically indistinguishable pathogens (molecular clusters), state the sampling fraction, the distribution of cluster sizes, and the study population turnover, if known | Cluster definitions, number and sizes described in results |
| Descriptive data | 14* | (a) Give characteristics of study participants (eg, demographic, clinical, social) and information on exposures and potential confounders  b) Indicate the number of participants with missing data for each variable of interest  (c) *Cohort study*—summarise follow-up time (eg, average and total amount) | STROME-ID 14.1: give information by strain type if appropriate, with use of standardised nomenclature | This paper has not focused on the characteristics of participants (such as demography), but at the higher level of whether they were HCWs or patients, and characteristics of the facilities in which they were working or admitted – these are given in Results and Table 1 |
| Outcome data | 15* | *Cohort study*—report numbers of outcome events or summary measures over time  *Case-control study*—report numbers in each exposure category, or summary measures of exposure  *Cross-sectional study*—report numbers of outcome events or summary measures | ·· | Included data were primarily descriptive/quantitative |
| Main results | 16 | (a) Give unadjusted estimates and, if applicable, confounder-adjusted estimates and their precision (eg, 95% conﬁdence interval). Make clear which confounders were adjusted for and why they were included (b) Report category boundaries when continuous variables were categorised   If relevant, consider translating estimates of relative risk into absolute risk for a meaningful time period | STROME-ID 16.1: consider showing molecular relatedness of strain types by means of a dendrogram or phylogenetic tree | We considered including a phylogenetic tree, but with multiple different overlapping investigations (due to a highly-clonal outbreak), it would not have been decipherable. |
| Other analyses | 17 | Report other analyses done (eg, analyses of subgroups and interactions, and sensitivity analyses) | ·· | N/A |
| **Discussion** |  |  |  |  |
| Key results | 18 | Summarise key results with reference to study objectives | ·· | Summarised in Discussion (first paragraph) |
| Limitations | 19 | Discuss limitations of the study, taking into account sources of potential bias or imprecision. Discuss both direction and magnitude of any potential bias | STROME-ID 19.1: consider alternative explanations for ﬁndings when transmission chains are being investigated, and report the consistency between molecular and epidemiological evidence | Included in Discussion  Sequences for community cases included in the analysis to identify likelihood of multiple independent introductions (and number of introductions to each facility stated as a minimum number of introductions) |
| Interpretation | 20 | Give a cautious overall interpretation of results considering objectives, limitations, multiplicity of analyses, results from similar studies, and other relevant evidence | ·· | Included in discussion |
| Generalisability | 21 | Discuss the generalisability (external validity) of the study results | ·· | Included in discussion |
| **Other information** |  |  |  |  |
| Funding | 22 | Give the source of funding and the role of the funders for the present study and, if applicable, for the original study on which the present article is based | ·· | Funders had no role in the study |
| Ethics | 23 | ·· | STROME-ID 23.1: report any ethical considerations with speciﬁc implications for infectious-disease molecular epidemiology | No specific ethical implications identified |
